# Deep brain stimulation does not modulate fMRI resting-state functional connectivity in essential tremor

**DOI:** 10.1101/2023.02.02.23285325

**Authors:** Amar Awad, Filip Grill, Patric Blomstedt, Lars Nyberg, Johan Eriksson

## Abstract

**Background:** While the effectiveness of deep brain stimulation (DBS) in alleviating essential tremor (ET) is well-established, the underlying mechanisms of the treatment are unclear. ET, as characterised by tremor during action, is proposed to be driven by a dysfunction in the cerebello-thalamo-cerebral circuit that is evident not only during motor actions but also during rest.

**Methods:** DBS effects on resting-state functional connectivity were investigated by functional MRI in 16 ET patients with fully implanted DBS in the caudal zona incerta during On and Off therapeutic stimulation, in a counterbalanced design. Functional connectivity was calculated between different constellations of sensorimotor as well as non-sensorimotor regions (as derived from seed-based and data-driven approaches), and compared between On and Off DBS.

**Results:** We found that DBS did not modulate resting-state functional connectivity in ET.

**Conclusions:** The lack of DBS modulation during resting-state, in combination with previously demonstrated effects on the cerebello-thalamo-cerebral circuit during motor tasks, suggest an action-dependent modulation of DBS.

## Introduction

While the effectiveness of deep brain stimulation (DBS) in alleviating essential tremor (ET) is well-established, the underlying mechanisms of the treatment remain unclear. ET, the most common movement disorder, is caused by a dysfunctional cerebello-thalamo-cerebral (CTC) circuit. This dysfunction results in pathological tremor oscillations during movement, but the neuronal activity within the circuit has also been reported to be distorted during rest, without evident tremor (Raethjen and Deuschl, 2012; Holtbernd and Shah, 2021). In the current study, we investigate the effects of DBS on the CTC circuit dynamics in ET patients during resting-state.

DBS in the posterior subthalamic area (PSA), including the caudal zona incerta (cZi), and the ventral intermediate thalamic nucleus (Vim) is effective in alleviating tremor (Blomstedt et al., 2009; Deuschl et al., 2011; Fytagoridis et al., 2012; Groppa et al., 2014; Barbe et al., 2018; Kvernmo et al., 2022). Although DBS is thought to exert its effect by interrupting the propagation of tremor oscillations at the level of the stimulated target (cZi or Vim) (Plaha et al., 2008; Blomstedt et al., 2009; Groppa et al., 2014), it has been shown to modulate the neuronal activity of the entire CTC circuit (Ceballos-Baumann et al., 2001; Perlmutter et al., 2002; Haslinger et al., 2003; Gibson et al., 2016; Awad et al., 2020). Indeed, by combining task-based fMRI with cZi/PSA-DBS during different motor tasks, we showed that DBS resulted in modulation of the sensorimotor CTC circuit BOLD signal in a complex manner as exhibited by task-depended as well as task-independent effects (Awad et al., 2020). Investigating DBS effects during motor tasks, with and without tremor as in our previous study, was motivated as tremor in ET is present during action and rarely during rest (Cohen et al., 2003; Bhatia et al., 2018).

Notably, functional imaging studies indicate abnormalities in the neuronal activity of the CTC circuit not only during motor tasks but also during rest, when the circuit is not engaged and tremor not present (for review see: Holtbernd and Shah, 2021; Pietracupa et al., 2021). ET pathophysiology has been examined in several resting-state fMRI (rs-fMRI) studies showing differences in functional connectivity within the CTC circuit as compared to healthy controls (Holtbernd and Shah, 2021; Lan et al., 2021). For example, functional connectivity has been shown to be decreased between the cerebellum and the sensorimotor cortex, decreased between primary and premotor sensorimotor cortices, and increased between the cerebellum and thalamus (Buijink et al., 2015; Lenka et al., 2017; Nicoletti et al., 2020; Tikoo et al., 2020). Furthermore, functional connectivity among regions outside the sensorimotor network, such as the default mode and frontoparietal networks, has also been reported to be altered in ET (Benito-León et al., 2015; Fang et al., 2015). Whether those alternations in functional connectivity are affected by DBS is still unknown and has not been studied before. Here, we aimed to study cZi-DBS effects on BOLD fluctuations as measured by rs-fMRI during On and Off therapeutic DBS in ET patients. We predicted that DBS would modulate the functional connectivity within the CTC circuit.

## Material and methods

### Patients and Surgical procedure

We included 16 patients with ET (9 male; average age 70 years, range 52-80 years) and fully-implanted DBS in the cZi/PSA. Out of 60 ET patients with cZi DBS at our department, 35 were excluded due to cognitive impairment/dementia, claustrophobia, significant head tremor, or MR-incompatible DBS systems. Of the remaining 25 patients, 17 consented but one died from unrelated causes before the initiation of the study. ET diagnosis was set by a movement-disorders specialist according to the ‘‘consensus statement of the Movement Disorder Society on Tremor’’ (Deuschl et al., 2008). A new consensus on the classification of tremor (Bhatia et al., 2018) was established after the diagnosis of our patients and the conduction of the study, but this was not deemed to change the diagnosis of the included patients. See Table 1 for patient demographics.

**Table 1:** Patient demographics and stimulation parameters.

| **Patient** | **Sex** | **Handed-**<br>**ness** | **Months**<br>**since**<br>**surgery** | **Family**<br>**history** | **Active DBS**<br>**lead during**<br>**fMRI** | **Stimulation parameters**<br>**(amplitude, pulse**<br>**width, frequency)** |
| --- | --- | --- | --- | --- | --- | --- |
| 1 | M | L | 27 | Yes | Right | 2.3 V, 60 μs, 130 Hz |
| 2 | F | R | 70 | No | Left | 2.7 V, 60 μs, 150Hz |
| 3 | F | R | 36 | Yes | Left | 1.2 V, 60 μs, 160 Hz |
| 4 | F | R | 54 | Yes | Left | 1.3 V, 60 μs, 140 Hz |
| 5 | F | R | 43 | Yes | Right | 1.5 V, 60 μs, 130 Hz |
| 6 | M | R | 17 | Yes | Left | 1.8 V, 60 μs, 160 Hz |
| 7 | M | R | 34 | Yes | Left | 1.6 V, 60 μs, 140 Hz |
| 8 | F | R | 37 | Yes | Left | 2.3 V, 60 μs, 140 Hz |
| 9 | M | R | 36 | No | Left | 1.8 V, 60 μs, 160 Hz |
| 10 | F | R | 11 | Uncertain | Left | 1.5 V, 60 μs, 140 Hz |
| 11 | M | R | 44 | Yes | Left | 1.6 V, 60 μs, 140 Hz |
| 12 | F | R | 59 | Yes | Left | 1.8 V, 60 μs, 150 Hz |
| 13 | M | R | 59 | No | Left | 2.2 V, 60 μs, 160 Hz |
| 14 | M | R | 26 | Yes | Left | 2.3 V, 60 μs, 160 Hz |
| 15 | M | R | 50 | Yes | Left | 2.5 V, 60 μs, 140 Hz |
| 16 | M | R | 17 | No | Left | 1.7 V, 60 μs, 140 Hz |

The patients had been receiving chronic DBS in the cZi/PSA with a stable clinical response for at least 1 year (range 1 – 5.8 years). The implantation of the electrodes was done under general anaesthesia without microelectrode recording or intraoperative test stimulation. The target in the cZi/PSA was visually identified on a stereotactic T2-weighted MRI slightly posteromedial to the posterior tip of the subthalamic nucleus at the level of the maximal diameter of the red nucleus (Fig. 1). The location of the electrodes was verified using an intraoperative, or postoperative, CT fused with the preoperative MRI. The patients were implanted with electrode model 3389 Medtronic and a single “implanted pulse generator” (Activa, Medtronic). All patients gave written informed consent, and the study was approved by the local medical ethical board and was performed in accordance with the Declaration of Helsinki.

**Figure 1:**
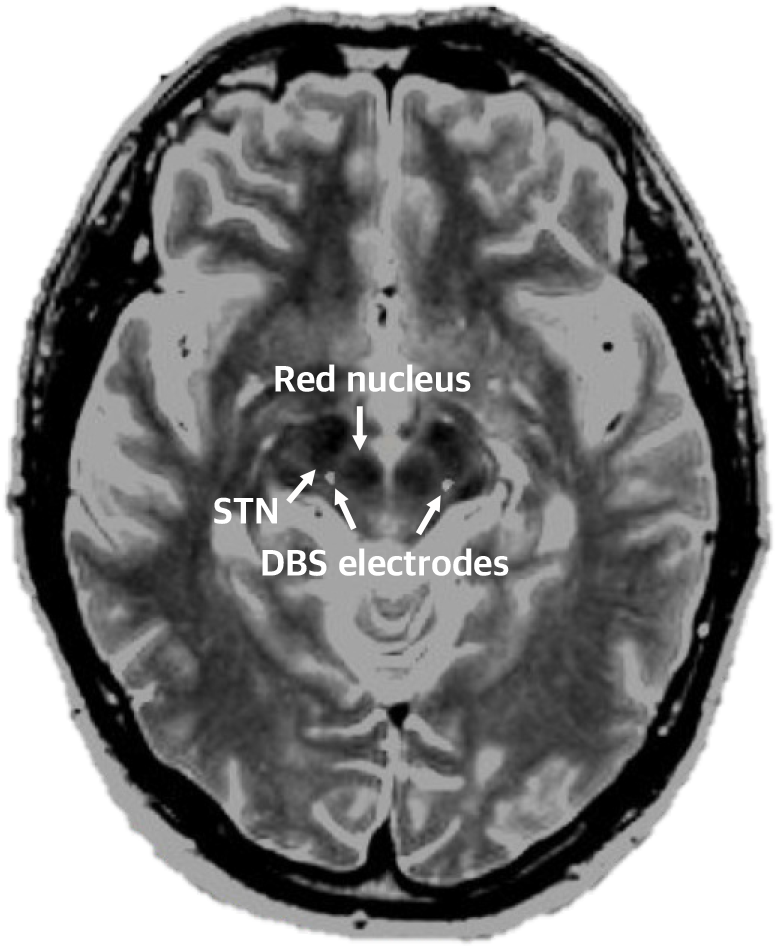
The DBS target (cZi/PSA). A preoperative axial MR-image fused with a postoperative CT for a representative patient, demonstrating the localisation of the tips of the DBS electrodes in the caudal zona incerta (cZi) within the posterior subthalamic area (PSA); posteromedial to the subthalamic nucleus (STN) at the level of the maximal diameter of the red nucleus.

### fMRI data acquisition and experimental design

All scans were performed with a Philips Achieva dStream 1.5 T MR scanner using a transmit-receive (T/R) head coil, and average head-specific absorption rate (SAR) below 0.1 W/kg. During each DBS condition (On and Off), three experiments/runs were collected: task-based fMRI with different motor tasks as previously published in Awad et al., 2020, rs-fMRI (current study), and task fMRI with a working memory task (unpublished). Functional echo-planar imaging (EPI) rs-fMRI runs were performed with the following parameters: 31 interleaved axial slices at a TR 3000 ms, TE 50 ms, flip angle 90°, voxel size 3.44 × 3.49 × 4.4 mm, 0.5 mm inter-slice gap, field of view (FOV) 220 × 220 mm, and matrix size 64 × 63. Axial T1-weighted structural scan was collected after the first functional session with the following acquisition parameters: 180 slices, no inter-slice gap, 1 × 1 x 1 mm voxel size, TR 7.4 s, TE 3.4 ms, flip angle 8°, FOV 256 × 232 mm, and matrix size 256 × 232.

Two rs-fMRI time-series were collected per patient, one for each stimulation condition (unilateral On and Off cZi-DBS). The first five volumes were discarded prior to each session to allow fMRI signal equilibrium. For each acquisition, 154 volumes (∼ 8 minutes) per session were collected. The patients were instructed to lie still in the scanner with their eyes opened and focusing on a fixation cross presented on a screen that was seen via a double-mirror mounted on the head coil.

Therapeutic unilateral left-sided DBS was used in all, except two, patients (who had right-sided DBS activated during the On session). For patients implanted with bilateral DBS electrodes (n= 5), the right electrode (ipsilateral to tested arm during the motor task-experiment) was switched off during the whole experiment. The initial stimulation setting was counterbalanced across patients, i.e. half of the patients started the first session with DBS Off and the other half with DBS On. therapeutic stimulation parameters were used during the study. These parameters were previously optimised for maximal tremor reduction without side effects.

The head movements were restricted by using foam padding between the head and head coil in all patients. To further restrict head movements, bite bars fixed on the head coil were used when tolerated (6 patients). These bite bars were custom-made for each patient to match the patient’s own teeth prior to the scanning session. Furthermore, foam padding was used between the head and head coil for additional head-motion restriction in all patients.

### Data analyses

#### Image processing

Image data (T1 and EPI volumes) from the two patients with active right-sided electrode during the On session were flipped with respect to the mid-sagittal plane prior to pre-processing.

##### Pre-processing of fMRI data

fMRI data were pre-processed using CONN toolbox version 20b based on SPM12 implemented in MATLAB. Images were realigned, unwarped, and slice-time corrected. Outlier volumes were detected using the Artifact Detection Tools (ART) as implemented in CONN and by using the option for conservative threshold; an image was defined as an outlier if the head displacement in the x, y, or z direction was greater than 0.5 mm from the previous frame, or if the global mean intensity of an image was greater than 3 SD from the mean image intensity for the entire resting scan. The images were then normalised to the standard MNI space and smoothed with an 8-mm full-width at half-maximum Gaussian kernel. Functional and structural T1-weithted images were segmented into grey matter, white matter, and CSF.

##### Denoising

fMRI data were further denoised by using component-based noise correction method (*CompCor*) implemented in CONN. Twelve realignment parameters and their quadratic effects (Friston24-parameters), potential outlier scans, and signal from white matter and cerebrospinal fluid masks were used as confounds. Further, the data were bandpass-filtered (0.008-0.09 Hz). Global signal regression was not applied.

A group-specific anatomical template was created from the individual 16 T1-weighted images using DARTEL (diffeomorphic anatomical registration through exponentiated lie algebra) for a more precise inter-subject alignment (Ashburner, 2007). A group-specific anatomical image was created by averaging individual normalised T1-weighted images and then used to create a group-specific rendered brain image.

#### fMRI data analyses

##### Identifying the sensorimotor network and creating sensorimotor regions of interest (ROIs)

The sensorimotor circuit was defined based on seed-to-voxel functional connectivity with the Yeo-17 left-motor-cortex ROI (Yeo et al., 2011). We chose this ROI because it generated a more inclusive/complete sensorimotor network (especially regarding the left hemisphere where DBS was turned On and Off during the experiment) as compared to other tested seeds, see supplementary figure 1. Across both rs-fMRI runs (during On and Off DBS) of each patient, the Pearson’s correlation between fMRI time series at each voxel and the left-motor-cortex ROI was computed. The resultant functional connectivity map is shown in figure 2 at p < 0.0001 uncorrected. Sensorimotor ROIs were created as spheres around relevant peak coordinates with a 5 mm radius, except for the supplementary motor area (SMA) where 10 mm was used to include both hemispheres. The ROIs included primary motor cortex, premotor cortex, SMA, postcentral gyrus, thalami, putamen, cerebellum lobule V/VI and cerebellum lobule VIII), see fig. 2 below

**Figure 2:**
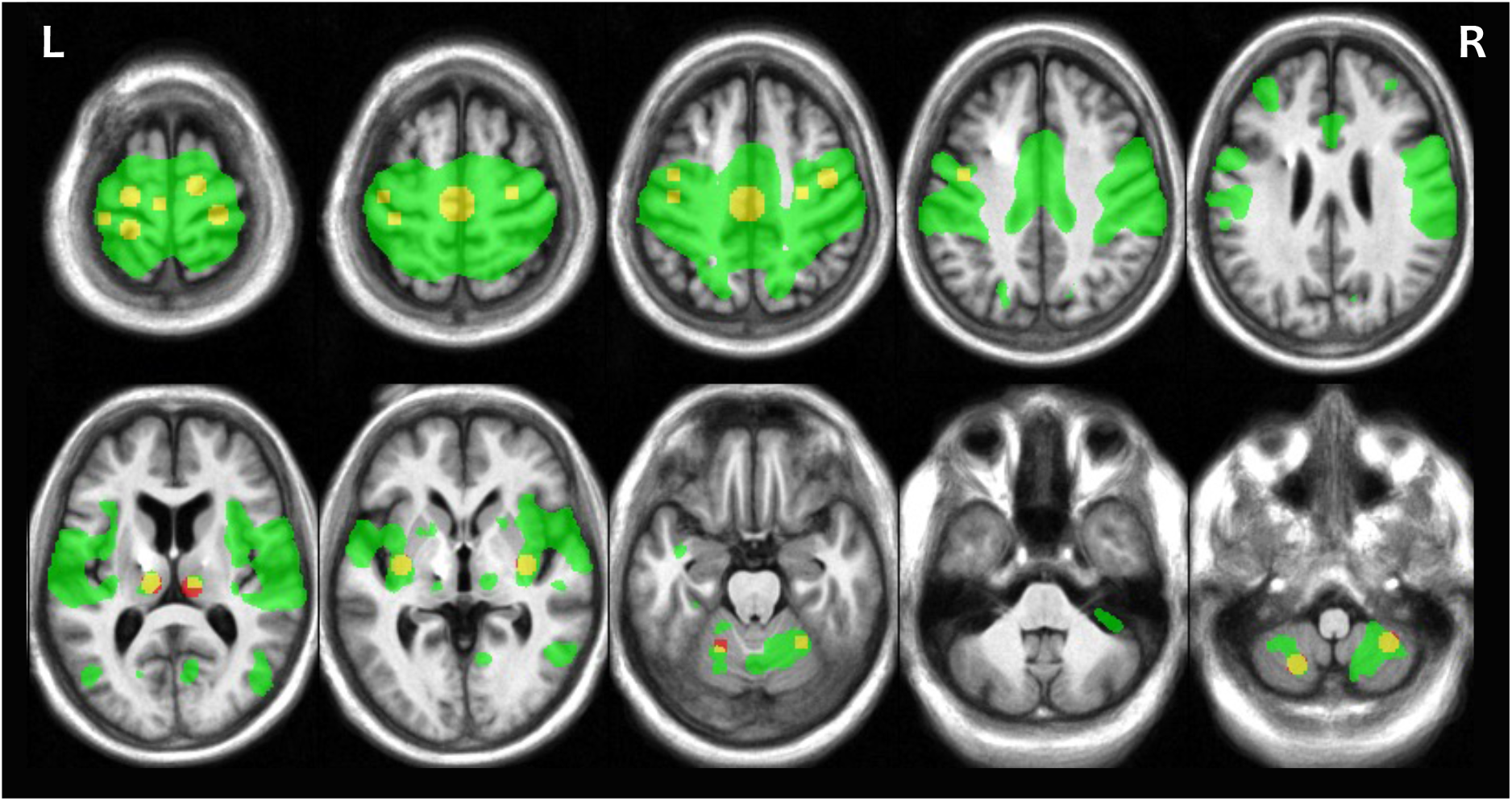
Sensorimotor network and ROIs. The sensorimotor functional connectivity map as extracted from voxel-wise correlation to Yeo-17 left-motor-cortex ROI (green), and the created sensorimotor ROIs (red/yellow).

The analyses of cZi-DBS effects on slow BOLD fluctuations as measured by rs-fMRI during On and Off therapeutic DBS in ET patients were done in 4 steps, as further detailed below.

##### Calculation of functional connectivity between averaged sensorimotor ROIs

To examine widespread and bilateral functional connectivity differences between the cerebral cortex, cerebellum, putamen and cerebellum, ROIs from these locations were grouped and treated as one ROI. That resulted in four grouped ROIs: cerebral cortex regions bilaterally, bilateral thalamic regions, bilateral putamen regions, and all cerebellar regions bilaterally; see fig. 3. Cross-correlation was computed between extracted average time-series from grouped ROIs during On and Off sessions. Correlation coefficients were then Fisher transformed to z values. Paired sample t-tests were used to calculated correlation value differences between On and Off DBS.

**Figure 3:**
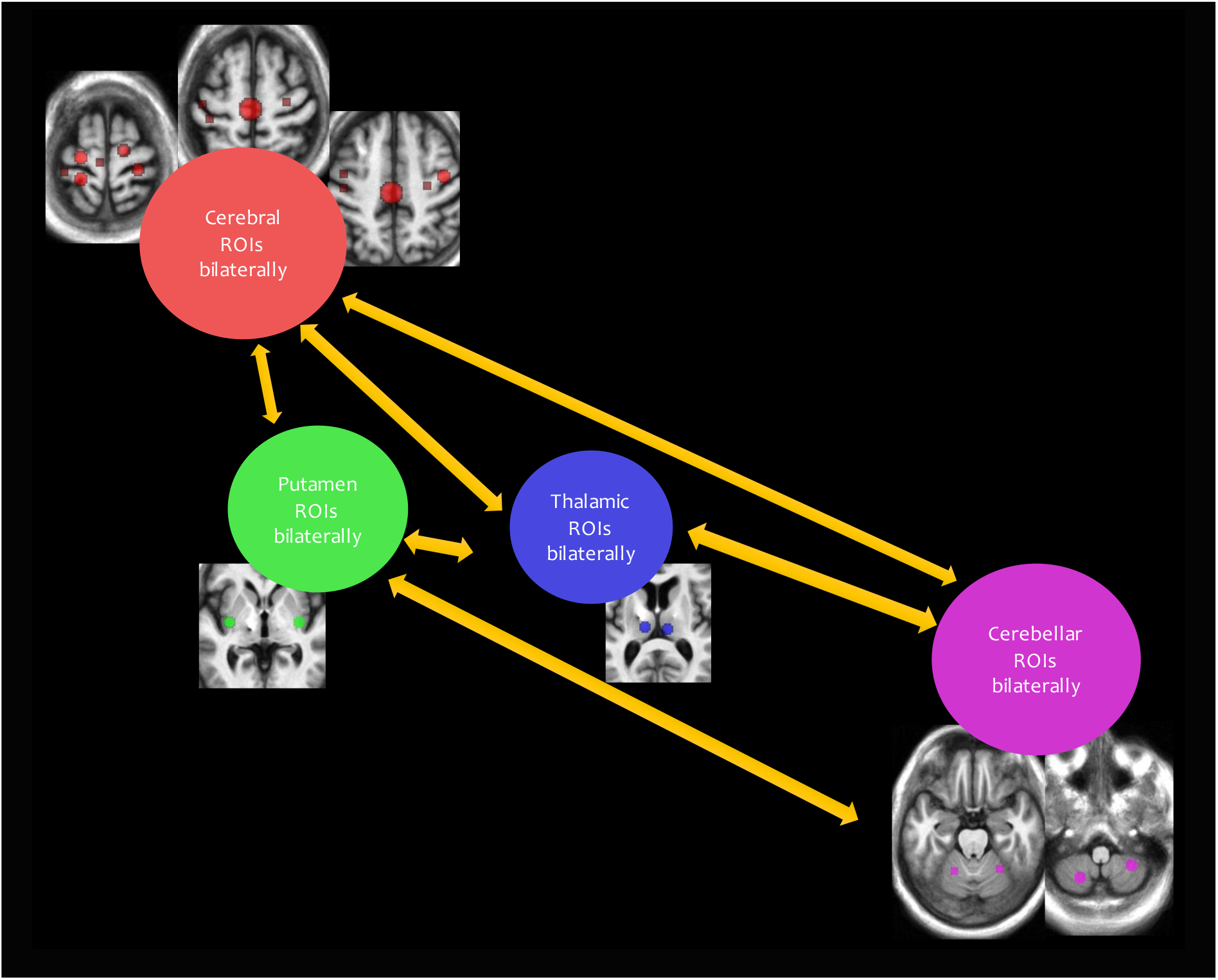
Calculation of functional connectivity between averaged sensorimotor ROIs. The four grouped sensorimotor ROIs and their six connections that were used to calculate wide-spread bilateral functional connectivity differences between On and Off DBS.

##### Calculation of functional connectivity between sensorimotor ROIs separately for each hemisphere

Cross-correlations were computed between time-series from each ROI in one cerebral hemisphere with time-series from cerebellar ROIs in the opposite side, which resulted in 21 tested connections for each hemisphere. Correlation coefficients were then Fisher transformed to z values. Paired sample t-tests were used to calculated correlation value differences between On and Off DBS. This analysis is more sensitive than the previous step since BOLD signal is not averaged across regions. It is motivated by the lateralised anatomical connections through crossing fibres between the cerebellum and cerebrum. Further, only unilateral DBS was active in the current study which prompted examination of each hemisphere separately.

##### Calculation of ALFF within sensorimotor ROIs (grouped and separated)

The power spectrum of each ROI was obtained by transforming the ROI timeseries to the frequency domain. The mean square root of the power in the frequency range across 0.01 – 0.1 Hz were used as a measure of ALFF (Zou et al., 2008). The ALFF values were calculated for grouped and separated ROIs, similar to the two previous steps in functional connectivity analysis. The procedure was performed on DBS Off and On fMRI runs separately. The ALFF score for each ROI and fMRI run was then entered into a paired t-test to investigate differences of ALFF as a function of DBS. Several rs-fMRI studies have shown altered ALFF values in the CTC circuit in ET patients (Gallea et al., 2015; Yin et al., 2016; Wang et al., 2018).

##### Dual-regression to investigate DBS effects on resting-state networks identified through independent component analysis (ICA)

Dual-regression ICA was conducted in 14 patients (excluding the two with active right-sided DBS during fMRI), and followed the steps suggested by Nickerson et al (Nickerson et al., 2017). Prior to denoising, fMRI runs from both Off and On sessions as well as for each individual were concatenated and entered into a group ICA restricted to 30 components. The resulting group average components were visually inspected and identified according to known resting-state networks: visual, frontoparietal, lateral sensorimotor, default mode, salience, medial sensorimotor, cerebellar and ventral attention network. The remaining 22 networks represented noise such as movement artefacts, BOLD signal from white matter and ventricles. Each group-average network was then regressed into each subject’s and condition’s time resolved dataset giving subject and condition specific time series. The network specific time series where then regressed into the same time resolved dataset yielding subject and condition specific functional connectivity maps. The resulting connectivity maps were then entered into a paired samples t-test using FSL’s randomise function (Winkler et al., 2014) to look for differences between DBS On and Off (5000 permutations, threshold-free cluster enhancement (TFCE) corrected). This step is motivated by rs-fMRI studies indicating functional connectivity abnormalities in resting-state networks beyond the sensorimotor circuit (Benito-León et al., 2015; Fang et al., 2015)

## Results

The average framewise displacements as calculated according to the approach suggested by (Power et al., 2012) did not differ between Off and On conditions (Off: 0.14 ± 0.09 mm, On: 0.16 ± 0.10 mm, p= 0.52, paired-sample t-test).

### DBS differences in functional connectivity between averaged sensorimotor ROIs

There was no statistically significant difference in correlation values between On and Off DBS when calculating the differences in functional connectivity between averaged/grouped bilateral sensorimotor ROIs (all p’s > 0.24 paired-sample t-test, uncorrected), see fig. 4.

**Figure 4:**
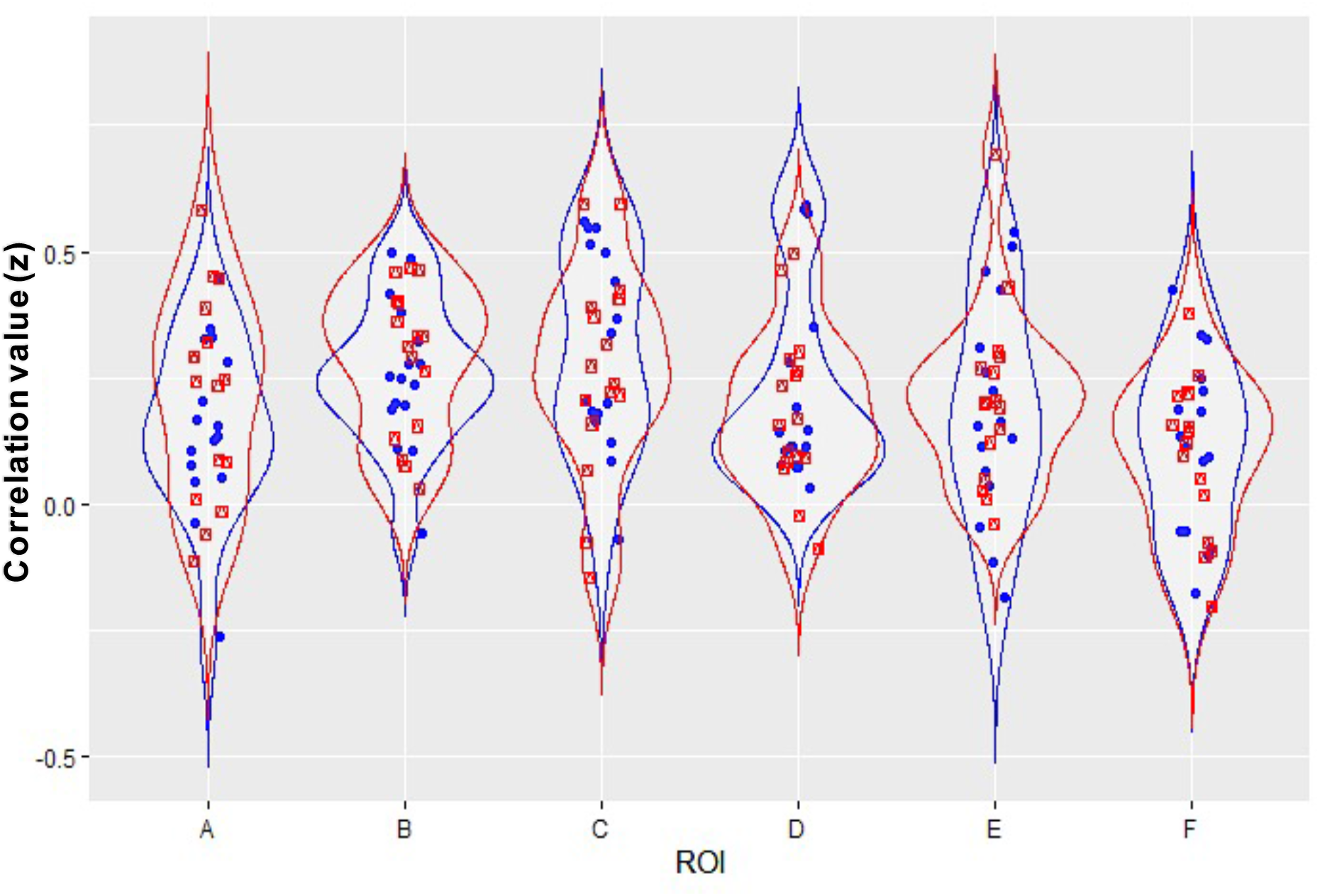
Functional connectivity between averaged sensorimotor ROIs. The diagram illustrated the relatively similar distributions in correlational values between On and Off DBS. Correlation values (z) are showed on the y-axis, and connections on the x-axis: A) thalamus-cerebral cortex, B) putamen-cerebral cortex, C) thalamus-putamen, D) cerebellum-cerebral cortex, E) cerebellum-thalamus, and F) cerebellum-putamen. Blue = Off DBS, red = On DBS.

### DBS differences in specific functional connectivity between sensorimotor ROIs separately for each hemisphere

No statistically significant difference was detected in correlation values between On and Off DBS when calculating differences in functional connectivity between separate sensorimotor ROIs within each hemisphere (all p > 0.09 paired-sample t-test, uncorrected), see supplementary fig. 2.

### DBS differences in ALFF within sensorimotor ROIs (averaged and separated)

The ALFF values in sensorimotor ROIs, both averaged and separated, did not differ significantly between On and Off DBS (all except one test with p > 0.13 paired-sample t-test, uncorrected). There was a difference for the ALFF value in the left dorsal premotor cortex ROI with p= 0.03, which until replicated in an independent sample, should be considered non-significant given many tests without correction for multiple comparisons.

### DBS differences in multiple resting-state network as calculated via dual-regression ICA

Using ICA, eight networks were identified as shown in fig. 5. Dual-regression analysis did not show a statistically significant difference between DBS On and Off (TFCE-corrected, p > .05) in any of the 8 identified components.

**Figure 5:**
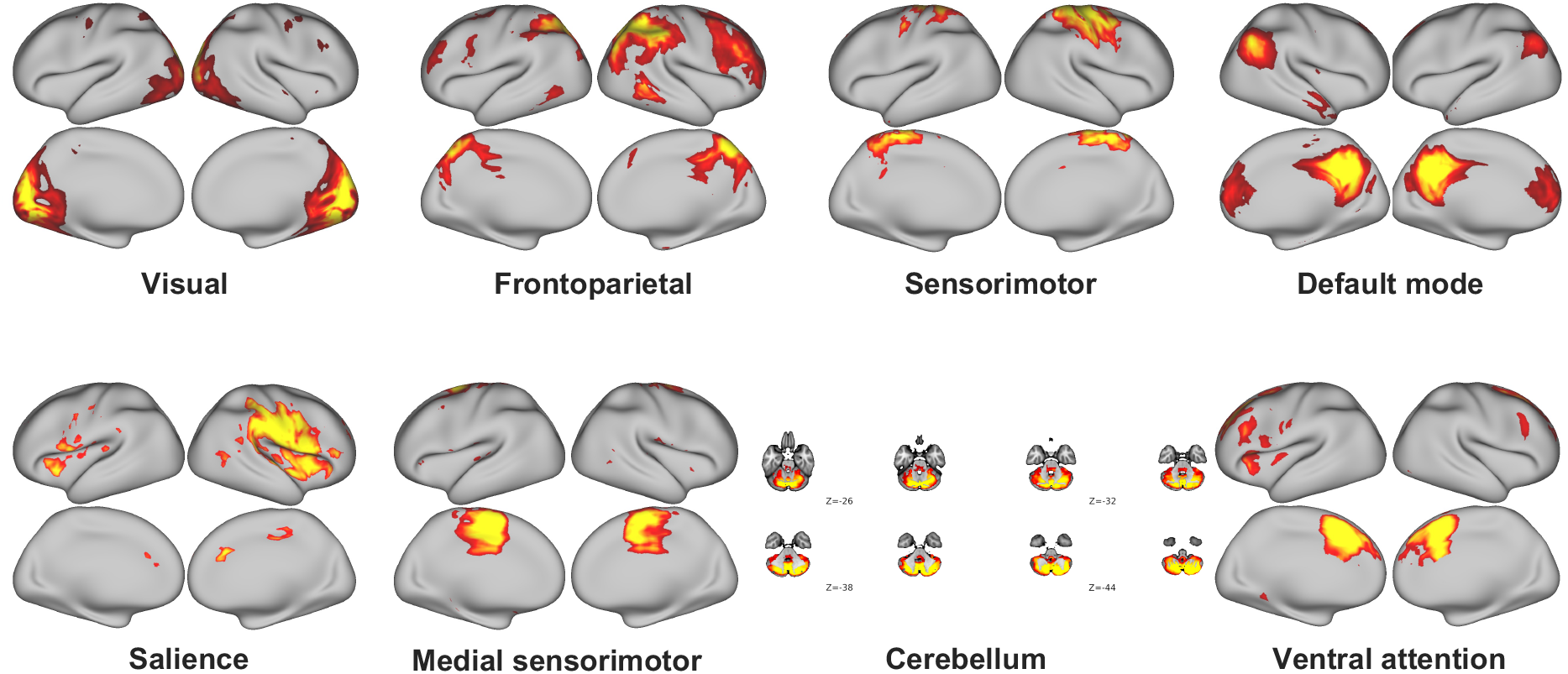
Resting-state networks as identified by ICA. 8 networks/components could be mapped to canonical resting-state networks. The remaining 22 components reflected noise (not shown here)

## Discussion

We investigated cZi-DBS effects on slow BOLD fluctuations as measured by rs-fMRI during On and Off therapeutic DBS in ET patients and found no significant modulation of resting-state functional connectivity. This was the case when examining DBS effects on i) widespread functional connectivity between sensorimotor cerebral cortex, thalamus, putamen, and cerebellum; ii) hemisphere-specific functional connectivity in ROIs within the aforementioned regions; iii) ALFF within sensorimotor ROIs; and iv) multiple well-known resting-state networks, sensorimotor as well as non-sensorimotor. Thus, the correlation in BOLD signal fluctuations among nodes, within and outside the cerebello-thalamo-cerebral circuit, is comparable in Off and On DBS.

Since this is the first study to examine the the effects of DBS on functional connectivity in ET, a comparison with other studies is difficult. However, the null-findings of this study could be compared to findings from previous reports about the effects of Vim-thalamotomy on functional connectivity. In a number of publications with overlapping populations and findings, Tuleasca et al investigated rs-fMRI functional connectivity in ET patients before (Tuleasca et al., 2018c, 2018a) and after (Tuleasca et al., 2018b, 2020) stereotactic radiosurgical thalamotomy with Gamma knife. They reported several rs-fMRI networks before or after unilateral thalamotomy to be correlated with tremor reduction postoperatively (Tuleasca et al., 2018c, 2018a, 2018b). However, they applied ICA for identification of resting-state networks and included all or most of the resultant networks, including some that might represent noise from motion or respiration artefacts, white matter and ventricles (Tuleasca et al., 2018c, 2018a). Also, the altered connectivity patterns reported by Tuleasca and colleagues were mainly based on correlations with tremor reductions, while we have here refrained from using correlations due to the small sample size (Marek et al., 2022). In summary, the methodological limitations of the aforementioned studies (Tuleasca et al., 2018c, 2018a, 2018b, 2020) limit further analyses of why thalamotomy, but not DBS, might affect the functional connectivity in ET.

Although being one of the largest DBS-fMRI studies, the sample size of the current study is still small, and the study might simply be underpowered to detect potential effects of interest. However, the distributions of ROI-ROI correlation values are relatively similar during On and Off DBS (fig. 4 and supplementary fig. 2), which imply that modest effects would still be hard to find even with a much larger sample size. Moreover, negative findings were demonstrated despite (deliberately) liberal statistical testing, and are thus unlikely to represent false negative findings.

The choice of ROIs was based on their functional specifications (Poldrack, 2007). They represented well-known nodes within the sensorimotor network (fig. 4). Therefore, we consider it unlikely that the choice of ROIs impacted the results negatively. Also, functional connectivity and ALFF were probed with different constellations of connections, from averaged-ROI-connections to capture potential widespread changes, to individual ROI-connections to capture potential specific changes between ROIs. Moreover, the dual-regression analysis was based on ICA, which is a data-driven method for identifying networks independent on the choice of ROIs (Stone, 2002; Nickerson et al., 2017).

MR-signal drop-out due to the DBS hardware is another potential limitation. The metallic objects in the DBS electrodes and extension cables are known to result in susceptibility artefacts (signal loss) mostly pronounced around the electrode tip and the left parietal cortex (Kahan et al., 2012; Gibson et al., 2016; Boutet et al., 2020). Obviously, this precluded the acquisition of useful image data from those areas which affected the lateral part of the sensorimotor network, and the salience network that seemed to be right-dominated. Importantly, most of the sensorimotor circuit (fig. 2) and other circuits (fig. 5) are not disturbed by signal drop-out. Due to safety concerns related to heating of the DBS-system when exposed to the radiofrequency pulses of the MR scanner, we adjusted the imaging parameters in order to keep the SAR values below 0.1 W/kg (Georgi et al., 2004; Carmichael et al., 2007; Kahan et al., 2015). For example, we used lower static magnetic field strength (1.5 Tesla), and a T/R head coil to reduce radiofrequency exposure to the DBS system. These compromises resulted in lower signal-to-noise ratio. Despite these limitations, our data were of sufficient quality to generate a well-known sensorimotor network based on ROI-to-voxel functional connectivity, and moreover, ICA could identify the canonical resting-state networks (fig. 2 and fig. 5).

The present finding that DBS did not affect resting-state functional connectivity within and outside the sensorimotor circuit can be related to our previous observation of DBS effects on functional brain activity (Awad et al., 2020). In that previous study, differences in BOLD-signal amplitude during DBS On versus Off were assessed for a postural holding task, a pointing task, and a resting control task. A main result was that DBS-On led to reduced activity in primary sensorimotor cortex and cerebellum (lobule VIII) during the postural task but not during rest. This observation is in good agreement with the present finding of no DBS effects during resting state. In addition, in Awad et al. (2020) it was found that DBS-On led to increased activity in left premotor cortex during all tasks (the postural and pointing tasks as well as rest), and even to selective increases in activity during the rest condition in the SMA and the cerebellum (lobule IV/V). One obvious explanation of why DBS effects were seen on functional brain activity at rest but not on resting-state functional connectivity concerns the different analytic approaches. Task effects capture transient modulation of blood flow and BOLD signal, whereas functional connectivity might reflect stable functional networks of regions that typically are co-activated and minimally influenced by brief interventions. By this view, modulation of the BOLD-signal amplitude during rest by DBS could reflect elements of motor preparedness/planning and task-set switching (i.e., getting prepared for the upcoming postural holding task and task set switching from the pointing task to rest) (Sakai, 2008; Baker et al., 2011) that are not taxed during a long period of rest in resting-state fMRI. Thus, the modulation of BOLD signal during rest as well as motor tasks in Awad et al. (2020) might reflect multiple aspects of action, and we therefore propose that DBS modulation of the sensorimotor circuit in ET is action-dependent in a broad sense. This notion is coherent with the fact that DBS alleviates tremor, which in ET is action tremor that is present during action and rarely during rest (Cohen et al., 2003; Bhatia et al., 2018).

## Conclusions

In this study, DBS effects on functional connectivity in ET were probed by rs-fMRI during On and Off therapeutic DBS in the cZi/PSA. DBS did not modulate resting-state functional connectivity in sensorimotor or non-sensorimotor networks. As DBS previously was shown to modulate the cerebello-cerebral circuit during motor tasks (Awad et al., 2020), we argue that DBS modulation is action-dependent.

## Data Availability

All data supporting the conclusions of this manuscript will be made
available by the authors upon request to any qualified researcher.

## Abbreviations

ALFF: amplitude of low frequency fluctuations
BOLD: blood oxygen level-dependent
cZi: caudal part of zona incerta
CTC: cerebello-thalamo-cerebral
DBS: deep brain stimulation
rs-fMRI: resting-state functional magnetic resonance imaging
PSA: posterior subthalamic area
SMA: supplementary motor area
Vim: ventral intermediate nucleus of the thalamus

## Acknowledgements

We would like to thank all the patients who have taken part in this study; AnnaKarin Kronhamn for recruitment of patients and assistance during the experiments; and Anders Lundquist for statistical support.

