## Supplementary material for "Deep brain stimulation does not modulate fMRI resting-state functional connectivity in essential tremor": -

### Appendix A: Supplementary figures

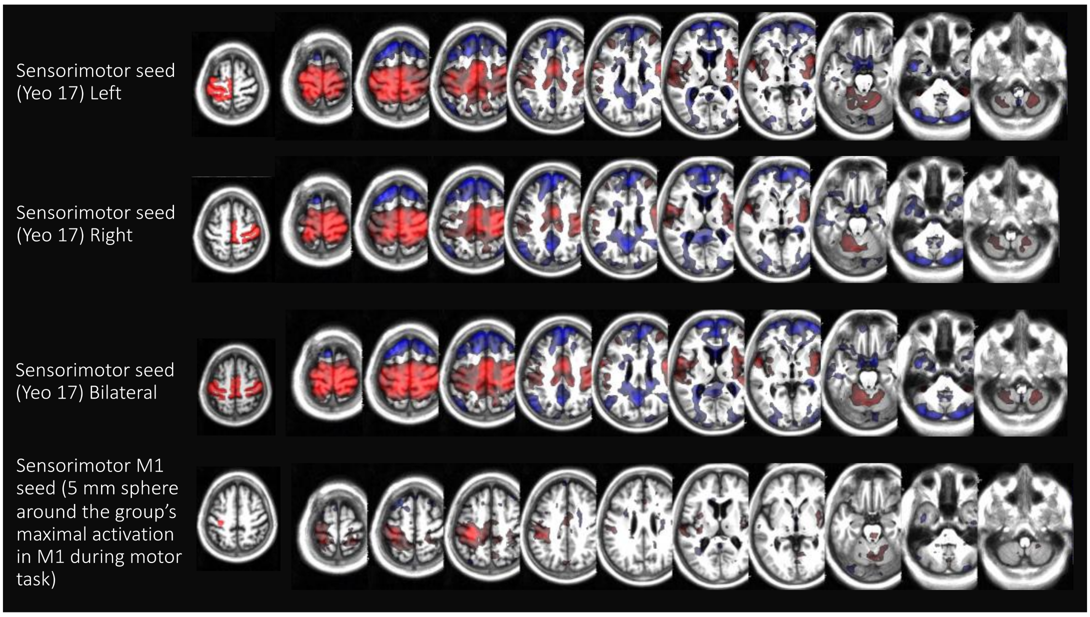


**Supplementary figure 1: Seed-to-voxel analysis to extract the sensorimotor network from four different seeds.** The generated sensorimotor maps were visually compared *ad hoc*. The left sensorimotor seed from Yeo-17 (uppermost panel) was chosen since it generated a complete sensorimotor network with all expected regions (cerebral, thalamic, basal ganglia and cerebellar), and was more sensitive for generating sensorimotor maps from the left cerebral (and right cerebellar) hemisphere that was modulated by DBS during the experiment.

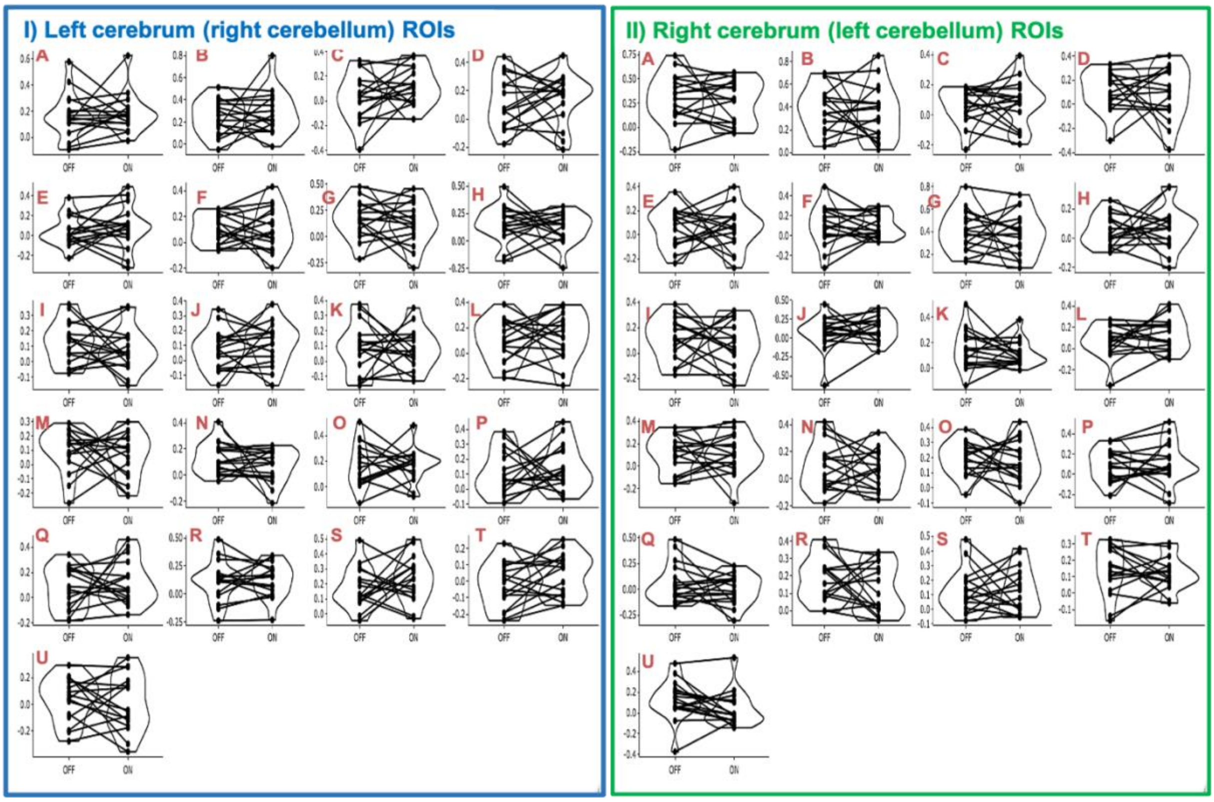


**Supplementary figure 2:** Functional connectivity between sensorimotor ROIs as separated by hemisphere, I) left cerebral and right cerebellar ROIs and II) right cerebral and left cerebellar ROIs. One plot for each connection and correlation values (z) are showed on the y-axes. Connections: A) M1-premotor, B) M1-SMA, C)M1-thalamus, D) M1-cerebellum V, E) M1-cerebellum VIII, F) M1-putamen, G) Premot-SMA, H) Premot-thalamus, I) Premot-cerebellum V, J) Premot-cerebellum VIII, K) Premot-putamen, L) SMA-thalamus, M) SMA-cerebellum V, N) SMA-cerebellum VIII, O) SMA-putamen, P) Thalamus-cerebellum V, Q) Thalamus-cerebellum VIII, R) Thalamus-putamen, S) cerebellum V-cerebellum VIII, T) cerebellum V-putamen, U) Cerebellum VIII-putamen. M1= primary motor cortex, SMA = supplementary motor area.
